# The association between disability and all-cause mortality in low and middle-income countries: a systematic review and meta-analysis

**DOI:** 10.1101/2023.03.21.23287520

**Authors:** Tracey Smythe, Hannah Kuper

**Affiliations:** International Centre for Evidence in Disability, London School of Hygiene & Tropical Medicine, Keppel st, WC1E7HT, London, UK; Division of Physiotherapy, Department of Health and Rehabilitation Sciences, Stellenbosch University, Cape Town, South Africa

## Abstract

**Background:** There are at least one billion people with disabilities globally. On average they have poorer health, yet worse healthcare access. We aimed to systematically review the association between disability and mortality in low- and middle-income countries (LMICs).

**Methods:** We searched MEDLINE, Global Health, PsycINFO and EMBASE from 1st January 1990 to 14th November 2022. We included any longitudinal epidemiological study in any language with a comparator group that measured the association between disability and all-cause mortality in people of any age. Two reviewers independently assessed study eligibility, extracted data, and assessed risk of bias. We used a random-effects meta-analysis to calculate the pooled hazard ratio (HR) for all-cause mortality by disability status. We then conducted meta-analyses separately for different impairment and age groups.

**Findings:** We identified 6146 unique articles, of which 70 studies (81 cohorts) were included in the systematic review, from 22 countries. There was variability in the methods used to assess and report disability, as well as mortality. The meta-analysis included 53 studies, representing 62 cohorts (comprising 267,415people with disabilities). Pooled HRs for all-cause mortality were 2.06 (95%CI 1.80 - 2.34) for people with disabilities versus those without disabilities, with high heterogeneity between studies (τ^²^=0·24, I^²^=98%). This association varied by impairment type; from 1.32 (95%CI 1.13 – 1.55) for visual impairment to 3.39 (95%CI 0.90 – 12.76) for multiple impairments. The association was highest for children under 18 (4.46, 95%CI 3.01–6.59); and lower in people aged 15 – 49 (3·53, 95%CI 1·29–9·66); and older people over 60 years (1·97, 95%CI 1·63–2.38).

**Conclusion:** Disability increases the risk of all-cause mortality in LMICs, particularly in childhood. Interventions are needed to improve health of people with disabilities and reduce their risk of death.

**Key messages:** *What is already known on this topic:* Globally, people with different impairments have a higher risk of death than those without disabilities and many deaths will be avoidable.

*What this study adds:* People with disabilities in LMIC have two-fold higher mortality rates that those without disabilities. Disability is associated with a higher hazard of age-adjusted all-cause mortality across diverse LMIC settings and populations as well as impairment types. The risk of dying early is highest for children with disabilities.

*How this study might affect research, practice or policy:* Improved understanding of the association between disability and mortality will help to inform public health planning and policy, and the allocation of limited health-care resources to optimise healthy longevity for all populations worldwide. Without a focus on disability it may be difficult to reach SDG3 and other key global health targets.

## Introduction

There are at least one billion people with disabilities globally, more than 80% of whom live in low and middle-income countries (LMICs) (1). People with disabilities include those who have long-term physical, mental, intellectual or sensory impairments which, in interaction with various barriers, may hinder their full and effective participation in society on an equal basis with others (2). Implicit in this conceptualisation are different pathways linking disability to premature mortality. People with disabilities, by definition, have an impairment and underlying health condition, which may increase their risk of death. For instance, people blind from diabetic retinopathy will still experience diabetes, which is linked to stroke and cancer (3, 4) . The impairment may also lead to further health conditions, linked to mortality (e.g. pressure sores or respiratory disease among people with physical impairment). Additionally, people with disabilities face a range of exclusions, so that they are more likely to live in marginalised and poor positions in society (5), which are linked to higher mortality (6). They also frequently face widespread difficulties and barriers in accessing healthcare, including inaccessible transport and facilities, poor skills of healthcare providers around disability, and high costs (7, 8). Consequently, people with disabilities have worse health than others in the population (9), are at higher risk of morbidity and mortality (1).

Many deaths will be avoidable. For instance, the confidential inquiry into premature deaths of people with intellectual disabilities in the UK estimated that 37% of deaths were avoidable from causes amenable to change by good quality healthcare (10). Data from the USA showed that people with disabilities were diagnosed with cancer at similar or earlier stages, but experienced higher rates of cancer-related mortality, implying health system failures in care (11). An improved understanding of the association between disability and mortality will help to inform public health planning and policy, and the allocation of limited health-care resources. Different systematic reviews have shown the relationship between specific impairment types (e.g. hearing (12), vision (13), intellectual (14, 15)) and mortality, but not disability overall. Furthermore, many of the studies included in these reviews are from High Income Settings, although 80% of people with disabilities live in LMICs. Age and impairment type are strong common risk factors for both disability and mortality (16, 17) and the association between disability and mortality may vary by these factors, yet this has not been explored to date. A comprehensive systematic review of disability and mortality in LMICs is therefore needed.

We undertook a systematic review and meta-analysis on the association between disability and age-adjusted all-cause mortality in all ages in LMIC. This systematic review and meta-analysis aimed to answer the following questions:

1. What is the extent, strength and quality of the published evidence of the association of disability and risk of all-cause mortality in LMIC?
2. To what degree does disability affect the risk of all-cause mortality, and does this risk vary based on age and impairment type in LMIC?

## Methods

### Search strategy and selection criteria

We developed a protocol for this systematic review and meta-analysis that was registered with Prospero (ref CRD42022302557) (18) and followed the Preferred Reporting Items for Systematic Reviews and Meta-Analyses guidelines (supplementary file 1). We systematically searched MEDLINE, Global Health, PsycINFO and EMBASE from 1st January 1990 to 14th November 2022 to identify available evidence in any language on disability and mortality in LMIC, including estimation of mortality rates and all-cause mortality.

Disability was conceptualised as arising from a combination of impairment and contextual barriers, as described in the following definitions: (i) UN Convention on the Rights of Persons with Disabilities (2006) (2) - People with disabilities include those who have long-term physical, mental, intellectual or sensory impairments which in interaction with various barriers may hinder their full and effective participation in society on an equal basis with others; (ii) World Report on Disability (2011) (1) - [Disability is] an umbrella term for impairments, activity limitations and participation restrictions, denoting the negative aspects of the interaction between an individual (with a health condition) and that individual’s contextual factors (environmental and personal factors). Search terms are outlined in supplementary file 2.

We included prospective and retrospective epidemiological studies (cohorts and randomised controlled trials) that measured the association of disability and all-cause mortality in any age. Conference abstracts were included if sufficient information was included for data extraction. There was no limitation of follow up period.

#### Inclusion criteria

1. Disability as defined in line with the International Classification of Functioning, Disability and Health Model and the United Nations Convention on the Rights of Persons with Disabilities (19). It includes people with specific conditions deemed likely to result in disability (e.g., spina bifida, schizophrenia), specific impairments (e.g. visual, hearing, physical) as well as disability measured through functioning/activity limitations/self-report (e.g., Washington Group questions, activities of daily living).
2. People of any age and sex, and any type of disability
3. LMIC, as defined by World Bank July 2021
4. Studies with a comparator group or that compared effect of disability on mortality to population statistics in a country as a whole (e.g. Standardised Mortality Ratio – SMR)
5. The outcome was all-cause mortality after baseline assessment of disability

Studies where all participants had a specific systemic disease (e.g. diabetes) were excluded due to issues with generalisability. Certain measures of disability were not eligible, including: 1) mild functional impairment (e.g. visual acuity ≥ 20/40, cognition as measured by MMSE ≥ 21, Washington group questions that reported ‘some difficulty’ or less, only one functional limitation/ limitation in activities of daily life (ADL)), 2) depression alone, 3) frailty alone, 4) disability as a continuous measure. Studies including both dementia and Alzheimer’s disease only had data extracted for dementia as Alzheimer’s was classified as a disease. Studies were excluded if there was no comparator group of people without disabilities or if they reported cause-specific mortality only (e.g. congenital heart disease, covid-19).

The outcome was all-cause mortality after baseline assessment of disability. Mortality could be reported using different measures of effect size and we included all measures. We also included cause of death and death rate.

### Data search and selection

The electronic search was conducted by TS. Results were screened and stored in Rayyan. Study selection was conducted independently by TS and HK. Each reviewer independently and systematically screened all titles and abstracts against the eligibility criteria. From these preliminary lists, the reviewers independently assessed the full text of each article for inclusion. The reason for exclusion was recorded and discrepancies discussed at each stage.

### Data extraction

To chart the data, we developed an extraction tool in Excel to systematically record information from included studies. The extraction form was piloted on four included studies to inform the final version. Each included study was charted by one reviewer and checked by the second reviewer.

Extracted information included:

1. Publication characteristics: author, title, year of publication, setting/country
2. Study design: study design, sample size
3. Participant characteristics: age, sex, disability measure, and any other relevant descriptive data
4. Outcomes: Effect size for mortality outcomes (e.g. hazard ratio, cumulative incidence ratio, death rate, odds ratio) and cause of death.

We extracted the crude estimates, and when estimates were reported with more than one level of adjustment, we extracted two additional estimates: (1) Minimally adjusted – usually the age and sex-adjusted estimate; and (2) Maximally adjusted - the greatest number of additional covariates, to elucidate potential pathways of the association of disability and mortality. When data on multiple cohorts were presented in a single publication, each cohort was separately eligible for inclusion. Where results from a study evaluated the association of disability and mortality over different population categories (e.g. non frail and frail people with disabilities) a weighted average of the effect measure was calculated for the entire cohort.

### Risk of bias assessment

The full texts of all eligible studies were assessed independently by both reviewers against quality assessment criteria adapted from Lund et al (20). The studies were evaluated for methodological quality and appropriateness for inclusion, without consideration of their results, based on a set of pre-determined criteria (Table 1). Disagreements on risk of bias ratings were resolved through discussion.

**Table 1:**
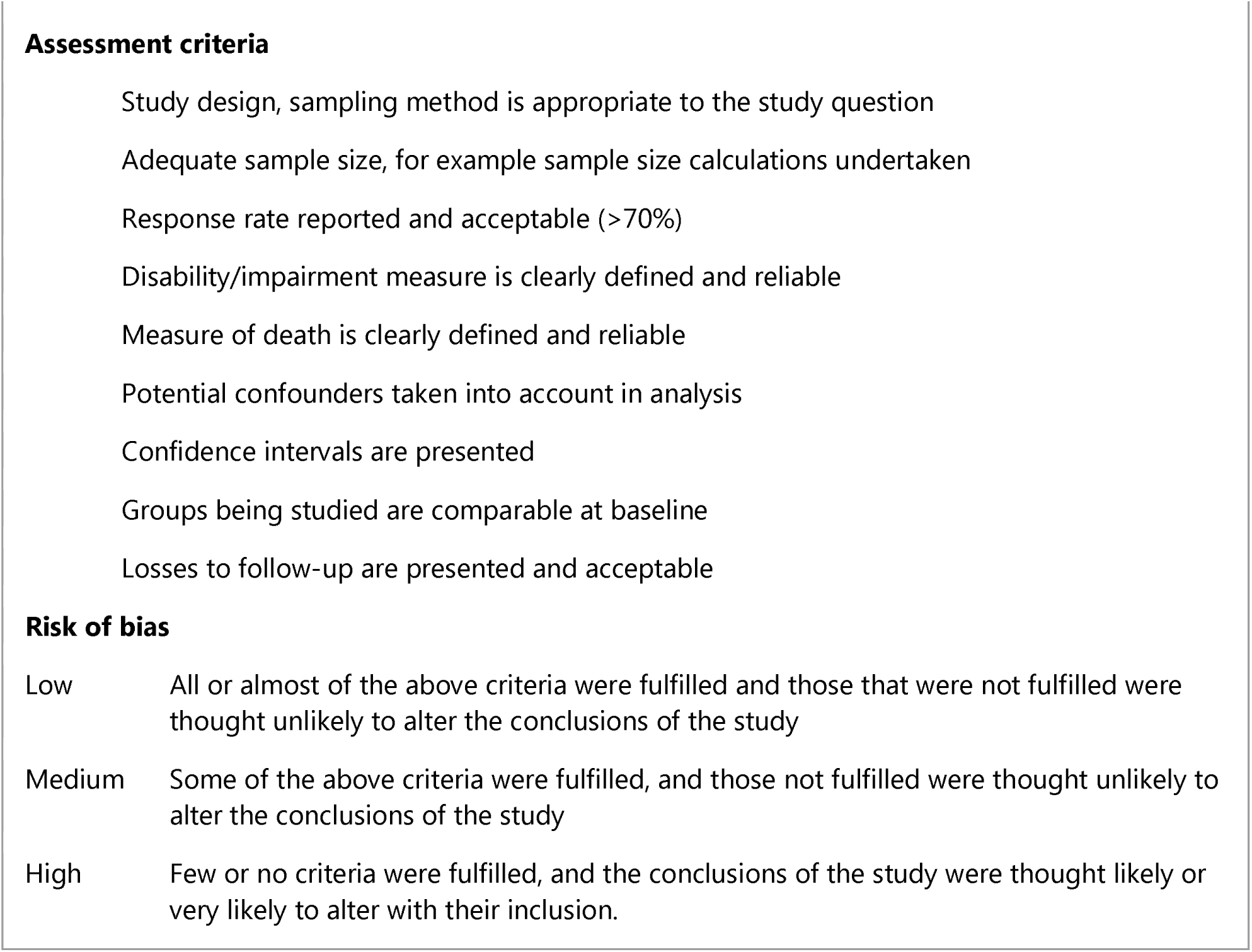
Risk of bias criteria and ratings

**Risk of bias**
|  |  |
| --- | --- |
| Low | All or almost of the above criteria were fulfilled and those that were not fulfilled were thought unlikely to alter the conclusions of the study |
| Medium | Some of the above criteria were fulfilled, and those not fulfilled were thought unlikely to alter the conclusions of the study |
| High | Few or no criteria were fulfilled, and the conclusions of the study were thought likely or very likely to alter with their inclusion. |

### Data Analysis

Meta-analyses were performed using a random-effects model to generate a pooled effect estimate, reported as the hazard ratio (HR) with 95% CIs for the association between disability and age-adjusted, all-cause mortality. Between-study heterogeneity was assessed with I^²^ and τ^²^ statistics. We included age-adjusted estimates of the effect of disability on mortality risk in the meta-analysis since age is a strong confounder for the association of mortality and disability. Where only multivariable adjusted estimates were available, these were used. Studies that did not adjust for age were therefore not included in the meta-analysis; studies that reported odds ratios were not included in the meta-analysis as the estimates could not be pooled with HRs. There is the potential for effect modification of the association by impairment type and age and therefore we undertook separate meta-analyses for different age groups (<=18 years, 15 years – 49 years and >=60 years) and impairment types (cognitive, functioning, hearing, neurological, multiple, physical, psychosocial and visual). We assessed the risk of publication bias using Egger’s test (threshold for significance p<0·05) and by inspection of funnel plots. A meta-regression was performed to estimate the between study variance and to calculate uncertainty in the estimated overall effect size risk of bias. We performed all statistical analyses using RevMan 5.4.1 (Cochrane collaboration).

Patient and public involvement

Patients were not involved in this research.

## Results

We identified 10,547 articles through electronic database searches. After removing 4,401 duplicate references, we screened the titles and abstracts of 6,146 articles. Of these, we identified 154 articles for full-text review. Eighty-four studies were excluded, leaving 70 studies that met the inclusion criteria (fig 1). Sixty-two cohorts from 53 studies were eligible for inclusion in the meta-analysis.

**Figure 1:**
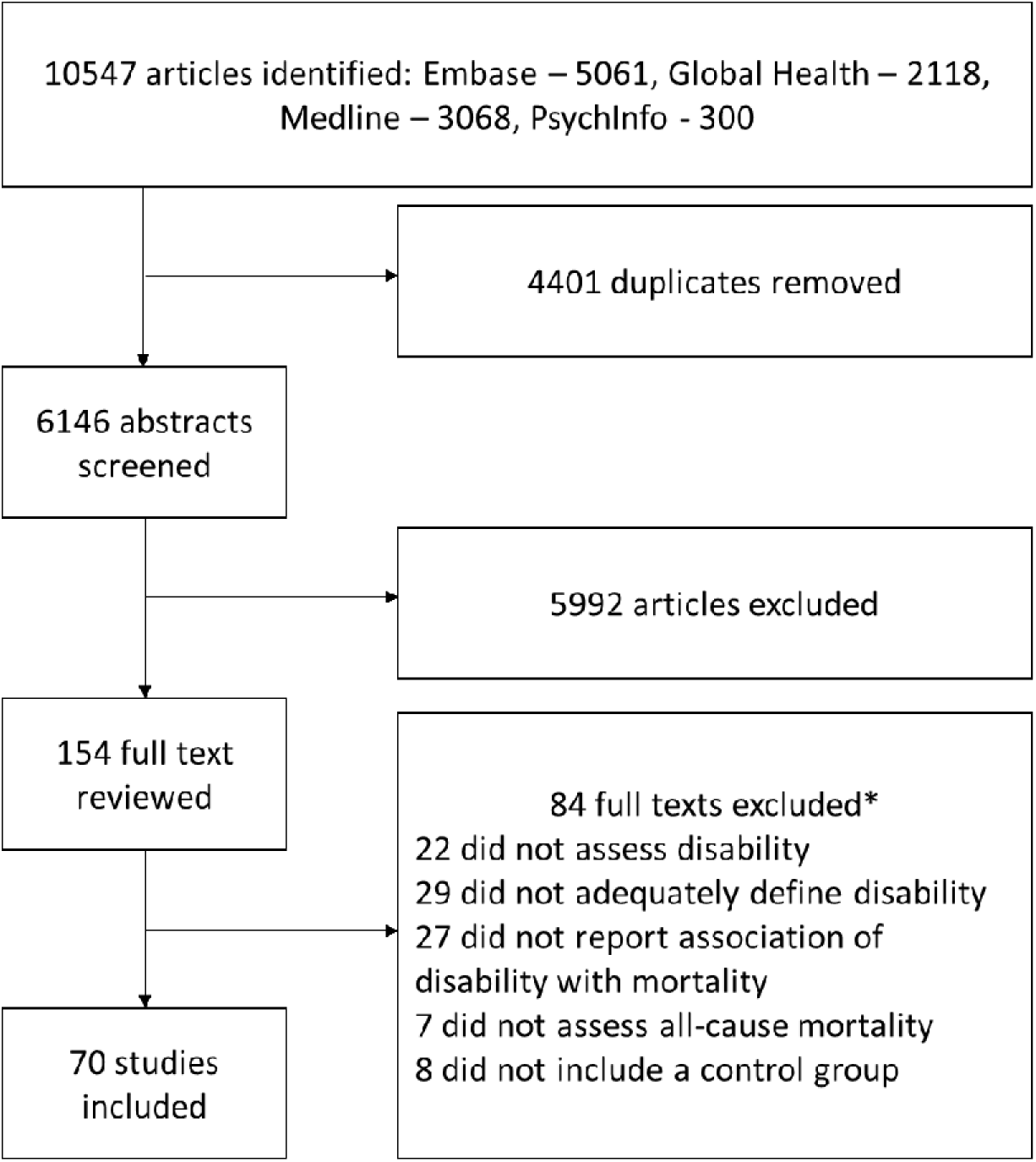
Study selection *some studies were excluded for more than one reason

The characteristics of the studies are reported in table 2. Geographically, the largest number of studies were conducted in the Western Pacific Region (n=33, 44%; China n= 32, Hong Kong n = 1) followed by the Region of the Americas (n=22, 29%; Brazil n=15, Cuba n=1, Dominican republic n=1, Mexico n=2, Peru n=2, Venezuela n=1), and the Africa Region (n=17, 22%; Cameroon n=1, Congo n=1, Ethiopia n=2, Kenya n=2, Malawi n=2, Nigeria n=2, South Africa n=1, Tanzania n=2, Togo n=1, Uganda n=1, Zimbabwe n=1, Zambia n=1). There were no studies from the Eastern Mediterranean region. The South-East Asian region was only represented by India (n=3), and the European region by Turkey (n=1). There is an increase in the number of studies collecting data on disability and death since 2010. The duration of follow up in included studies ranged from 28 days to 17 years. Sample sizes ranged from 14 (people with neurological impairment in China) (21) to 749,720 (people with neurological impairment in Brazil)(22).

**Table 2.**
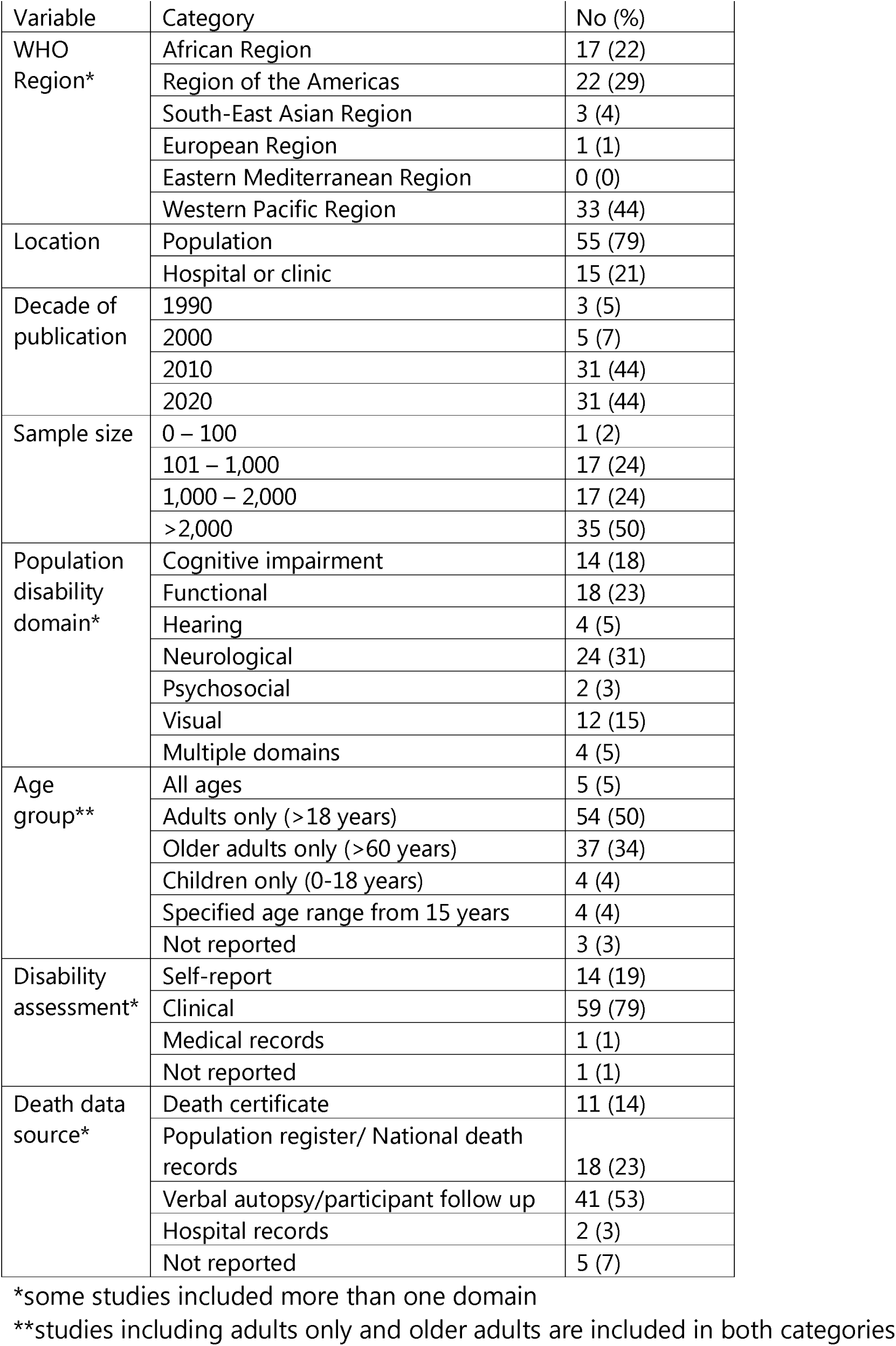
Study characteristics

| Variable | Category | No (%) |
| --- | --- | --- |
| WHO Region* | African Region | 17 (22) |
|  | Region of the Americas | 22 (29) |
|  | South-East Asian Region | 3 (4) |
|  | European Region | 1 (1) |
|  | Eastern Mediterranean Region | 0 (0) |
|  | Western Pacific Region | 33 (44) |
| Location | Population | 55 (79) |
|  | Hospital or clinic | 15 (21) |
| Decade of publication | 1990 | 3 (5) |
|  | 2000 | 5 (7) |
|  | 2010 | 31 (44) |
|  | 2020 | 31 (44) |
| Sample size | 0 – 100 | 1 (2) |
|  | 101 – 1,000 | 17 (24) |
|  | 1,000 – 2,000 | 17 (24) |
|  | >2,000 | 35 (50) |
| Population disability domain* | Cognitive impairment | 14 (18) |
|  | Functional | 18 (23) |
|  | Hearing | 4 (5) |
|  | Neurological | 24 (31) |
|  | Psychosocial | 2 (3) |
|  | Visual | 12 (15) |
|  | Multiple domains | 4 (5) |
| Age group** | All ages | 5 (5) |
|  | Adults only (>18 years) | 54 (50) |
|  | Older adults only (>60 years) | 37 (34) |
|  | Children only (0-18 years) | 4 (4) |
|  | Specified age range from 15 years | 4 (4) |
|  | Not reported | 3 (3) |
| Disability assessment* | Self-report | 14 (19) |
|  | Clinical | 59 (79) |
|  | Medical records | 1 (1) |
|  | Not reported | 1 (1) |
| Death data source* | Death certificate | 11 (14) |
|  | Population register/ National death records | 18 (23) |
|  | Verbal autopsy/participant follow up | 41 (53) |
|  | Hospital records | 2 (3) |
|  | Not reported | 5 (7) |

\*some studies included more than one domain
\*\*studies including adults only and older adults are included in both categories

Studies used a wide variety of measures to assess disability (Table 3). Fifty-nine studies (62 cohorts) used clinical assessments and 14 studies (19 cohorts) used self-report to measure disability. The most commonly used assessment for visual impairment was visual acuity testing with a standardised chart (23), and the mini-mental state examination (MMSE) questionnaire (24) was most commonly used to determine cognitive impairment. The Katz Index of Independence in Activities of Daily Living (ADL) (25) was the tool used most to evaluate functioning.

**Table 3:** Summary of included cohorts

| Primary author, year | Country (Study) | Sample size of people with disabilities | Female n% (participants with disabilities) | Comparator group sample size | Age | Setting | Baseline assessment period | Follow up duration | Disability assessment | Mortality assessment method |
| --- | --- | --- | --- | --- | --- | --- | --- | --- | --- | --- |
| **Cognitive** |  |  |  |  |  |  |  |  |  |  |
| An, 2018 (27) | China (Beijing longitudinal study of ageing) | 193 | 151 (81.9%) | 321 | 55+ | Population | 1992 | 8 years (median) | Clinical | Death certificate |
| Aprahamian, 2018 (28) | Brazil | 296 | 201 (67.9%) | 405 | 60+ | Hospital | 2014 - 2016 | 12 months (total) | Medical records and clinical | verbal autopsy |
| Ascencio, 2022a (29) | Peru | 53 | not reported | 374 | 60+ | Population | 2013 | 46.5 months | clinical | national health database |
| Campos, 2020 (30) | Brazil (Bambui Health Aging Study) | 21 | 12 (57.1%) | 1134 | 60+ | Population | 1997 | 11 years (mean) | Clinical | population register |
| Cao, 2022 (31) | China (China health and retirement longitudinal study (CHARLS)) | 1348 | not reported | 2581 | 60+ | population | 2011-2012 | not reported | clinical | participant follow up |
| Correa, 2021b (32) | Brazil (EpiFloripa Aging Cohort) | 330 | not reported | 1061 | 60+ | Population | 2009-2010 | 4 years | Clinical | population register |
| Duan, 2020 (33) | China (Chinese Longitudinal Healthy Longevity Survey (CLHLS)) | 7573 | not reported | 15896 | 65+ | Population | 1998 (waves 2002 and 2008) | 6 years | Clinical | verbal autopsy |
| Flaherty, 2011b (34) | China | not reported | not reported | not reported | 90+ | Population | not reported | 4 years | Self report | not reported |
| Gao, 2014 (35) | China | 400 | 297 (74.3%) | 1600 | 65+ | Population | 2003 - 2005 | 7 years | Clinical | verbal autopsy |
| Han, 2021 (36) | China (CLHLS) | 1882 | not reported | 6411 | 65+ | Population | 1998 (2011 - 2018 waves) | 7 years | Clinical | death certificate and verbal autopsy |
| Hao, 2018 (37) | China (Longevity and Aging in Dujiangyan (PLAD)) | 523 | 397 (75.9%) | 86 | 90+ | Population | 2005 | 4 years | Clinical | population register |
| Su, 2021 (38) | China (CLHLS) | 252 | Not reported | Not reported | 60+ | Population | 2009 - 2010 | 10.8 years (median) | Clinical | verbal autopsy |
| Wu, Z 2020 (39) | China (CHARLS) | 820 | 598 (72.9%) | 4293 | 60+ | Population | 2011 | 4 years | Clinical | not reported |
| Xavier, 2010 (40) | Brazil (Epidemiologia do Idoso) | 56 | not reported | 1,477 | 65+ | Population | 1991-1992 | 10 years | Clinical | verbal autopsy and death certificate |
| **Functioning** |  |  |  |  |  |  |  |  |  |  |
| Bahat, 2015 (41) | Turkey | not reported | not reported | Not reported | 60+ | Hospital | 1999 - 2010 | 40.4 months | Clinical | population register |
| Bento, 2021 (42) | Brazil (SABE study) | 177 | 0 (0%) | 686 | 60+ men | Population | 2000 | 14 years max | Clinical | population register |
| Cabrero Castro, 2021 (43) | Mexico (MHAS) | 1,614 | not reported | 10,661 | 50+ | Population | 2001 | 3 years | Clinical | verbal autopsy |
| Confortin, 2020 (44) | Brazil (EpiFloripa Aging Cohort) | 536 | 1087 (64%) | 457 | 60+ | Population | 2009-2010 | 4 years | Clinical | population register |
| Correa, 2021a (32) | Brazil (EpiFloripa Aging Cohort) | 426 | not reported | 965 | 60+ | Population | 2009-2010 | 4 years | Self report and clinical | population register |
| Flaherty, 2011a (34) | China | not reported | not reported | not reported | 90+ | Population | not reported | 4 years | Self report | not reported |
| Feng, 2010 (45) | China (CLHLS) | 817 | not reported | 12640 | 65+ | Population | 1998 (2002 wave) | 3 years | Self report and clinical | death certificate |
| Gbeasor-Komlanvi, 2020 (46) | Togo (Study of Hospitalised Older Adults) | 144 | not reported | 189 | 50+ | Hospital | 2018-2019 | 3 months | Clinical | verbal autopsy |
| Gray, 2016 (47) | Tanzania | 78 | not reported | Not reported | 70+ | Population | 2009-2010 | 3 years | Clinical | verbal autopsy |
| Hu, Z 2022 (48) | China (CHARLS) | 1,630 | 962 (59%) | 9,383 | 50+ | population | 2011 | 7 years | clinical | participant follow up |
| Kou, S 2022 (49) | China (CHARLS) | 3,791 | 2,882 (76%) | 33,382 | 60+ | population | 1998-2014 | 2-4 years | clinical | participant follow up |
| Lima-Costa, M 2011 (50) | Brazil (Bambui Cohort Study of Aging) | 28 | 22 (78.6%) | 1196 | 60+ | Population | 1997 | 8.8 | Self report | population register |
| Lyu, YB 2017 (51) | China (CLHLS) | Not reported | Not reported |  | ≥80 | Population | 2009 | 5 years | Clinical | verbal autopsy |
| Nascimento, 2018b (52) | Brazil (Bambui cohort study of aging) | IADL: 295 | IADL: 211 (81.5%) | IADL: 746 | 60+ | Population | 1997 | 15 year | Clinical | verbal autopsy and death certificate |
| Nascimento, 2018c (52) | Brazil (Bambui cohort study of aging) | ADL: 121 | BADL: 77 (63.6%) | 1060 | 60+ | Population | 1997 | 15 year | Clinical | verbal autopsy and death certificate |
| Prynn, 2020 (53) | Malawi (Karonga Health and Demographic Surveillance Site (HDSS)) | 1,277 | 857 (67.1%) | 15,471 | 15+ | Population | 2014-2015 | 29 months | Self report | verbal autopsy |
| Souza, 2021 (54) | Brazil (COMO VAI) | 125 | not reported | 920 | 60+ | Population | 2014 | 2.5 years | Self report | death certificate and population register |
| Tang, 2014 (55) | China (Beijing longitudinal study of ageing) | 74 | not reported | 1936 | 55+ | Population | 1992 | 17 years | Self report | verbal autopsy |
| Wu X, 2004 (56) | China (Beijing Multidimensional longitudinal study of aging ) | 481 | Not reported | 2377 | 55+ | Population | 2000 | 8 years | Self report | death certificate |
| **Hearing** |  |  |  |  |  |  |  |  |  |  |
| Agrawal, 2011b (57) | India | 210 | not reported | 1212 | 60+ | Population | 2008 | median: 518 days (3 days - 567 days) | Clinical | population register |
| Ascencio, 2022b (29) | Peru | 180 | not reported | 247 | 60+ | population | 2013 | 46.5 months | self report | national health database |
| Ojagbemi, 2017c (58) | Nigeria (Ibadan Study of Ageing) |  | not reported |  | 65+ | Population | 2003-2004 | 5 years | Self report | verbal autopsy |
| Sun, 2020b (59) | China (CLHLS) | 5277 | 3103 (58.8%) | 18884 | 65+ | Population | 1998 |  | Self report | not reported |
| **Multiple** |  |  |  |  |  |  |  |  |  |  |
| Lelijveld, 2020 (60) | Malawi (Pronut cohort) | 60 | 22 (36.7%) | 878 | 6-15.9 | Hospital | 2006 - 2007 | 7 years | Self report | verbal autopsy |
| Paixao, 2022 (61) | Brazil | 3308 | 1855 (53.2%) | 11477907 | children <3 years | hospital | 2015 - 2018 | 3 years | clinical | national health database |
| Sun, 2020c (59) | China (CLHLS) | 7774 | 5812 (74.8%) | 18884 | 65+ | Population | 1998 |  | Self report | not reported |
| Zheng, X 2015 (62) | China | 21,574 | not reported | national mortality rates | not reported | Population | 2006 | 4 years | Not reported | not reported |
| **Neurological** |  |  |  |  |  |  |  |  |  |  |
| Abuga, 2019 (63) | Kenya (Kilifi DHS) | 306 | 139 (48.9%) | 9912 survey controls<br>22873 age matched | children 6 - 9 | Population | 2001 | 14.7 years (median) | Clinical | verbal autopsy |
| Avelino, 2017 (64) | Brazil | 549 | 126 (69%) | 570 | 60+ | Hospital | 2009 - 2015 | 1 year max | Clinical | verbal autopsy |
| Bwakura-Dangarembizi, 2021 (65) | Zimbabwe and Zambia (HOPE-SAM) | 30 | not reported | 619 | <5 years | Hospital | 2016 - 2018 | 1 year | Clinical | verbal autopsy |
| Hong, 2005 (66) | China (CLHLS) | 161 | not reported | 3811 | ≥55 | Population | 2001 | 40 months | Clinical | Population register and verbal autopsy |
| Hu L, 2022 (67) | China | 72,102 | 37493 (52%) | population | all ages | hospital | 2011 | 8 years | clinical | death certificate |
| Jotheeswaran, 2010 (68) | India (10/66 Dementia Research Group) | 54 | 33 (61%) | 698 | 65+ | Population | 2004 - 2006 | 3 years | Clinical | verbal autopsy |
| Katzman, 1994 (69) | China | 53 | not reported | 3,351 | 65+ | Population | 1987 | 5 years | Clinical | verbal autopsy |
| Leite, 2019 (70) | Brazil | 434 | not reported | Population of Sao Paulo | All | Hospital | 2004 - 2014 | 4.8 years | Clinical | death certificate |
| Li, G 1991 (21) | China | 14 | not reported | 1076 | 60+ | Population | 1986 | 3 years | Clinical | verbal autopsy |
| Liu, T 2014 (71) | China (Second China National Sample Survey on Disability) | 2,071 | 1149 (55.5%) |  | 18+ | Population | 2006 | 4 years | Clinical | verbal autopsy |
| Melo, 2022 (22) | Brazil | 749,720 | 378,609 (50.50%) | 71,272,198 | all ages | hospital | 2000 - 2015 | 3 months - 15 years | clinical | national health database |
| Namaganda, 2020 (72) | Uganda | 97 | not reported | 41319 | 2-17 years | Population | 2005 | 54 months | Clinical | verbal autopsy |
| Niehaus, 2020 (73) | South Africa | subgroup of 60+; n=73 (from total of 981) | 30 (41.1%) |  | 60+ | Hospital | 1997-2005 | 12.1 years | Clinical | verbal autopsy |
| Nitrini, 2005 (74) | Brazil | 113 | 80 (70.8%) | 1280 | 65+ | Population | 1997 | 3 years | Clinical | population register |
| Ojagbemi, 2017a (58) | Nigeria (Ibadan Study of Ageing) | not reported | not reported |  | 65+ | Population | 2003-2004 | 5 years | Clinical | verbal autopsy |
| Ojagbemi, 2016 (75) | Nigeria (Ibadan Study of Ageing) | 255 | not reported | 1894 | 65+ | Population | 2003 - 2004 | 5 years | Clinical | verbal autopsy |
| Prince, 2012 (76) | Cuba, Dominican republic, Venezuela, Peru, Mexico, China (10/66 Dementia research group) | 775 | 538 (69.4%) | 11718 | 65+ | Population | 2003-2007 | 3-5 years | Clinical | verbal autopsy |
| Samba, 2016 (77) | Congo (EPIDEMCA-FU) | 63 | 49 (77.8%) | 791 | 65+ | Population | 2011 - 2012 | 2 years | Clinical | verbal autopsy |
| Teffera S, 2011 (78) | Ethiopia | 307 | 55 (17.9%) | national mortality rates | 15 - 49 | Population | not reported | 5 years | Clinical | verbal autopsy |
| Wang, 2019 (79) | China | 132 | not reported | Population | not reported | Hospital | 2007 | 10 years | Clinical | not reported |
| Wen, H 2011 (80) | China | 304 | not reported | 3581 | 55+ | Population | 1997 | 14 years | Clinical | verbal autopsy and certificate |
| Yung, 2022 (81) | Hong Kong | 8826 | 4687 (53.10%) | population | 18 - 39 | hospital | 2006 - 2012 | 5 years | clinical | national health database |
| Zhang, Y 2020 (82) | China | 132 | 75 (56.8%) |  | not reported | Clinic | 2007 | 10 years | Clinical | verbal autopsy |
| Zou, S 2022 (83) | China (China Kadoorie Biobank study) | 3080 | not reported | 254733 | 30-79 | population | 2004-2008 | 10 years | self report | population register |
| **Physical** |  |  |  |  |  |  |  |  |  |  |
| Nascimento, 2018a (52) | Brazil (Bambui cohort study of aging) | 348 | 239 (68.7%) | 765 | 60+ | Population | 1997 | 15 years | Clinical | verbal autopsy and death certificate |
| Nascimento, 2022 (84) | Brazil (SABE: (Health, Well-being and Aging Study) | 255 | not reported | 1156 | 60+ | population | 2000 | 10 years | clinical | national health database |
| **Psychosocial** |  |  |  |  |  |  |  |  |  |  |
| Fekadu, 2015 (85) | Ethiopia (Butajira cohort) | 919 | 347 (37.8%) | national mortality rates (2009) | 15 - 49 | Population | 1998 - 2001 | 11.3 years (median) | Clinical | verbal autopsy |
| Li, Y 2021 (86) | China | 112576 | 53488 (47.5%) |  | 15+ | Hospital | 2009-1014 | 5 years max | Clinical | population register |
| **Visual** |  |  |  |  |  |  |  |  |  |  |
| Agrawal, 2011a (57) | India | 134 | not reported | 128 | 60+ | Population | 2008 | median: 518 days (3 days - 567 days) | Clinical | population register |
| Ascencio, 2022c (29) | Peru | 321 | not reported | 106 | 60+ | population | 2013 | 46.5 months | self report | national health database |
| Cao, 2021 (87) | China | 218 | not reported | 4,942 | 30+ | Population | 2006 - 2007 | 6 years | Clinical | verbal autopsy |
| Gu, 2013 (88) | China (CLHLS) | 3,156 | not reported | 7,884 | 65+ | Population | 1998 (2002 - 2005 waves) | 3 years | Clinical | death certificate and verbal autopsy |
| Khanna, 2013 (89) | India (Andhra Pradesh Eye Disease Study) | 206 | not reported | 2991 | 30+ | Population | 1996 | 11 | Clinical | verbal autopsy |
| Kuper, 2019 (90) | Kenya (Nakuru Posterior Segment Eye Disease Study) | 341 | 159 (46.6%) | 3,100 | >50 | Population | 2007 - 2008 | 5.6 years | Clinical | verbal autopsy |
| Li Z, 2011 (91) | China (The Southern Harbin Eye Study) | Blind: 106<br>Moderate VI: 651 | Blind: 74 (69.8%)<br>Moderate VI: 408 (62.7%) | 4,300 | 50-96 | Population | 2005-2006 | 4 years | Clinical | verbal autopsy |
| Ojagbemi, 2017b (58) | Nigeria (Ibadan Study of Ageing) | 407 | not reported | 481 | 65+ | Population | 2003-2004 | 5 years | Self report | verbal autopsy |
| Pion, 2002 (92) | Cameroon | 101 | not reported | 101 | 40+ | Population | 1991 | 10 years | Clinical | verbal autopsy |
| Sun, 2020a (59) | China (CLHLS) | 5141 | 3344 (65%) | 18884 | 65+ | Population | 1998 |  | Self report | not reported |
| Taylor, 1991 (93) | Tanzania | 47 | 27 (57.4%) | 84 | All ages | Population | 1986 | 42 months | Clinical | verbal autopsy |
| Wang Z, 2022 (94) | China | 3644 | 2,219 (60.90%) | 12164 | 45+ | population | 2011 | 7 years | self report | participant follow up |

Studies used various strategies to assess mortality, and were included regardless of the methods used because official death registries might not have been available or provided high-quality data in many LMIC (26). Most cohorts (n=41) were followed up by methods such as verbal autopsy, interviews with key informants, or both, with some studies (n=18) searching official vital records. Table 3 summarises the characteristics of included studies.

All 70 studies (comprising 81 cohorts) calculated the association of mortality in participants with disabilities compared to those without disabilities (Table 4). There was high variability in effect estimates used for the association of disability and mortality, including HR (38 studies, 46 cohorts), OR (7 studies, 8 cohorts), RR (12 studies, 15 cohorts) and SMR (12 studies, 12 cohorts). Studies produced a combination of estimates; unadjusted (34 studies, 40 cohorts), age-sex adjusted (38 studies, 43 cohorts) and multivariable adjusted (40 studies, 47 cohorts). For risk of bias assessment (20), fifty-one (73%) studies (59 cohorts) received an assessment of low risk of bias across all nine domains. Six (9%) studies (8 cohorts) were assessed as having a high risk of bias, meaning that few criteria were fulfilled and the conclusions of the study are likely to alter with their inclusion.

**Table 4:** Association of disability with mortality

| Primary author, publication year | Age (years) | Deaths disability (n) | Deaths control (n) | Analysis | Unadjusted Mortality rate ratio (95%CI) | Age sex adjusted ratio (95%CI) | Multivariable adjusted ratio (95%CI) | Risk of bias |
| --- | --- | --- | --- | --- | --- | --- | --- | --- |
| **Cognitive** |  |  |  |  |  |  |  |  |
| An, 2018 | 55+ | 130 | 156 | Hazard ratio | 3.73 (2.95-4.71) | 2.77 (2.14-3.58) | 2.14 (1.59 - 2.87) | Low |
| Aprahamian, 2018 | 60+ | Not reported | Not reported | Hazard ratio |  |  | 3.86 (2.32 - 6.44) | Low |
| Ascencio, 2022a | 60+ | 18 | not reported | hazard ratio | 2.12 (1.28 - 3.51) |  |  | High |
| Campos, 2020 | 60+ | 17 | 538 | Hazard ratio | 2.5 (1.5-4.1) |  | 1.7 (1.0-2.7) | Low |
| Cao, 2022 | 60+ | not reported | not reported | Odds ratio |  | 1.59 (1.29 - 1.96) | 1.35 (1.08 - 1.69) | Low |
| Correa, 2021b | 60+ | Not reported | Not reported | Hazard ratio | 0.89 (0.63 - 1.24) |  | 0.82 (0.57 - 1.20) | Low |
| Duan, 2020 | 65+ | 3,523 | 3,360 | Hazard ratio | 3.25 (3.10–3.41) | 1.65 (1.56 -1.74) | 1.32 (1.25 - 1.41) | Low |
| Flaherty, 2011b | 90+ | Not reported | Not reported | Odds ratio |  |  | 1.54 (1.18 - 2.00) | Moderate |
| Gao, 2014 | 65+ | Not reported | Not reported | Hazard ratio | 1.48 (1.13-1.93) |  |  | High |
| Han, 2021 | 65+ | 1,560 | 2,652 | Hazard ratio | 3.88 (3.58 - 4.20) |  | 1.80 (1.64 - 1.96) | Low |
| Hao, 2018 | 90+ | 298 | 35 | Hazard ratio | 1.31 (0.88 - 1.94) |  | 1.75 (1.18 - 2.58) | Low |
| Su, 2021 | 60+ | Not reported | Not reported | Hazard ratio | 11.56 (5.91-22.61) | 3.83 (1.91-7.66) | 2.29 (1.24-5.04) | Low |
| Wu, Z 2020 | 60+ | 81 | 353 | Odds ratio |  | 1.6 (1.2-2.1) | 1.5 (1.1-2.0) | Low |
| Xavier, 2010 | 65+ | 44 | 930 | Relative risk | 2.69 (2.27 - 3.19) | 2.03 (1.66-2.47) | 1.64 (1.30 - 2.06) | Low |
| **Functioning** |  |  |  |  |  |  |  |  |
| Bahat, 2015 | 60+ | Not reported | 108 | Hazard ratio |  |  | 1.22 (1.12-1.32) | Moderate |
| Bento, 2021 | 60+ men | 25 | 38 | Hazard ratio | 2.5 (1.6-4.1) | 2.33 (1.89 - 2.95) | 1.96 (1.56 - 2.44) | Low |
| Cabrero Castro, 2021 | 50+ | 229 | 510 | Hazard ratio | 3.0 (2.6-3.4) |  |  | High |
| Confortin, 2020 | 60+ | 68 | 14 | Hazard ratio | 5.48 (2.31-12.99) |  | 2.97 (1.10-8.04) | Low |
| Correa, 2021a | 60+ | Not reported | Not reported | Hazard ratio | 1.32 (0.95 - 1.85) |  | 0.96 (0.65 - 1.4) | Low |
| Flaherty, 2011a | 90+ | Not reported | Not reported | Odds ratio |  |  | 1.41 (1.11 - 1.79) | Moderate |
| Feng, 2010 | 65+ | Not reported | Not reported | Hazard ratio |  |  | 1.9 (1.7-2.1) | Low |
| Gbeasor-Komlanvi, 2020 | 50+ | Not reported | Not reported | Hazard ratio |  |  | 2.49 (1.48 - 4.21) | Low |
| Gray, 2016 | 70+ | Not reported | Not reported | Odds ratio | 10.36 (7.35 - 14.60) |  | 2.67 (1.36 - 5.23) | Low |
| Hu, Z 2022 | 50+ | 778 | 408 | hazard ratio |  | 2.06 (1.81 - 2.34) | 1.63 (1.41 - 1.89) | Low |
| Kou, S 2022 | 60+ | not reported | not reported | risk ratio | 2.58 (2.52 - 2.65) | 2.07 (2.02 - 2.13) | 1.92 (1.86-1.97) | Low |
| Lima-Costa, M 2011 | 60+ | Not reported | Not reported | Hazard ratio |  |  | 3.4 (2.2-5.1) | Low |
| Lyu, YB 2017 | ≥80 |  | 571 | Hazard ratio |  | 1.37 (1.11 - 1.70) |  | Moderate |
| Nascimento, 2018b | 60+ | 216 | 314 | Hazard ratio | 1.7 (1.6-1.9) | 2.05 (1.70 - 2.48) |  | Moderate |
| Nascimento, 2018c | 60+ | 90 | 494 | Hazard ratio | 1.6 (1.4-1.8) | 1.92 (1.52 - 2.41) |  | Moderate |
| Prynn, 2020 | 15+ | 115 | 206 | Hazard ratio |  | 2.70 (1.91 - 3.82) | 2.65 (1.84 to 3.8) | Low |
| Souza, 2021 | 60+ | 54 | 58 | Risk ratio | 8.9 (6.1-12.9) |  | 3.1 (1.7-5.7) | Low |
| Tang, 2014 | 55+ | 58 | 1,010 | Hazard ratio | 1.5 (1.3-1.7) |  |  | Moderate |
| Wu X, 2004 | 55+ | Not reported | Not reported | Odds Ratio |  | 2.23 (1.53 - 3.23) |  | Moderate |
| **Hearing** |  |  |  |  |  |  |  |  |
| Agrawal, 2011b | 60+ | 28 | 72 | Hazard ratio | 2.4 (1.6-3.7). | 1.32 (0.8-2.19) | 1.22 (0.73-2.03) | Low |
| Ascencio, 2022b | 60+ | 41 | not reported | hazard ratio | 1.43 (0.93 - 2.21) |  |  | high |
| Ojagbemi, 2017c | 65+ | Not reported | Not reported | Risk ratio |  |  | 1.7 (0.8-2.2) | Low |
| Sun, 2020b | 65+ | 4678 | 9128 | Hazard ratio |  | 1.4 (1.3-1.4) | 1.3 (1.2-1.3) | Low |
| **Multiple** |  |  |  |  |  |  |  |  |
| Lelijveld, 2020 | 6-15.9 | 42 | Not reported | Risk ratio | 3.03 (1.65 - 5.56) |  | 6.99 (3.49 to 14.02) | Low |
| Paixao, 2022 | children <3 years | 398 | 120,629 | Mortality rate ratio | 14.3 (12.4 - 16.4) |  |  | Moderate |
| Sun, 2020c | 65+ | 9478 | 9,128 | Hazard ratio |  | 1.8 (1.7-1.8) | 1.5 (1.4-1.5) | Low |
| Zheng, X 2015 | not reported | 3,833 |  | SMR |  | 5.4 (5.2–5.5) |  | High |
| **Neurological** |  |  |  |  |  |  |  |  |
| Abuga, 2019 | children 6 - 9 | 11 | 92 survey 272 general population | Hazard ratio | 3.83 (2.05-7.16) | 4.24 ( 2.26-7.94) |  | Low |
| Avelino, 2017 | 60+ | 273 | 128 | Hazard ratio | 1.69 (1.02-2.80) |  | 1.29 (0.72 - 2.30) | Low |
| Bwakura-Dangarembizi, 2021 | <5 years | 10 | 45 | Hazard ratio | 5.12 (2.58-10.16) |  | 5.60 (2.72-11.50) | Low |
| Hong, 2005 | ≥55 | 69 | 266 | Relative risk |  | 1.63 ( 1.42 - 1.86) |  | Low |
| Hu L, 2022 | all ages | 11,766 | not reported | SMR |  | 10 - 39 years: 5.49 (5.34 - 5.36)<br>40 - 59 years: 4.01 (3.88 - 4.13)<br>60+ years: 4.85 (4.72 - 4.99)<br>All ages: 4.75 (4.00 - 5.63) |  | Low |
| Jotheeswaran, 2010 | 65+ | 25 | 210 | Hazard ratio | 2.8 (1.8 -4.3) | 2.3 (1.5 -3.7) | 2.4 (1.2-4.8). | Low |
| Katzman, 1994a | 65 - 74yr | 41 | 676 | Risk ratio |  |  | 7.19 (3.6 - 14.4) | Low |
| Katzman, 1994b | 75+ | 41 | 676 | Risk ratio |  |  | 3.49 (2.40 - 5.07) | Low |
| Leite, 2019 | All | 77 | Not reported | SMR |  | 2.88 (2.28-3.60) |  | Low |
| Li, G 1991 | 60+ | 8 | 40 | relative risk | 2.95 (not reported) |  |  | High |
| Liu, T 2014 | 18+ | 146 | Not reported | SMR by gender |  | men = 10.17 (not reported)<br>women = 12.42 (not reported) |  | Moderate |
| Melo, 2022 | all ages | 67,485 | 5,102,055 | relative risk | Overall: 1.27 (1.27 - 1.28)<br>male: 1.16 (1.32 - 1.35)<br>female: 1.34 (1.32 - 1.35)<br>15 - 29 yrs:2.82 (2.76 - 2.88)<br>30 - 59yrs: 1.42 (1.41 - 1.44) |  |  | Low |
| Namaganda, 2020 | 2-17 years | 15 | 169 | Hazard ratio |  | 2.90 (1.71-4.91) |  | Low |
| Niehaus, 2020 | 60+ | 23 (loss to follow up n=16) |  | Observed/ expected deaths |  | observed/ expected deaths = 1.77 (age adjusted) |  | Moderate |
| Nitrini, 2005 | 65+ | 58 | 163 | Hazard ratio | 5.16 (3.74–7.12) | 3.92 (2.80-5.48) |  | Low |
| Ojagbemi, 2017a | 65+ | Not reported | Not reported | Relative risk ratio |  |  | 3.5 (2.3-5.3) | Low |
| Ojagbemi, 2016 | 65+ | 137 | 698 | Hazard ratio | 1.8 (1.4-2.4) |  | 1.5 (1.1-2.1) | Low |
| Prince, 2012 | 65+ | 470 | 1681 | Hazard ratio |  | 2.77 (2.47 -3.10) |  | Low |
| Samba, 2016 | 65+ | Not reported | Not reported | Hazard ratio | 3.9 (2.4-6.3) |  | 2.5 (1.4-4.5) | Low |
| Teffera S, 2011 | 15 - 49 | 38 | Not reported | SMR |  | 5.98 (4.09-7.87) |  | Low |
| Wang, 2019 | not reported | 65 | Not reported | SMR |  | 1.2 (0.9-1.6) |  | Moderate |
| Wen, H 2011 | 55+ | 112 | Not reported | SMR |  | 0.91 (0.73-1.08) |  | Low |
| Yung, 2022 | 18 - 39 | 249 | Not reported | SMR |  | 1.23 (1.08 – 1.38) |  | Low |
| Zhang, Y 2020 | Not reported | 67 | Not reported | SMR |  | 1.23 (0.94–1.56) |  | Low |
| Zou, S 2022 | 30-79 | 97 | not reported | hazard ratio |  |  | 1.07 (0.87 - 1.30) | Low |
| **Physical** |  |  |  |  |  |  |  |  |
| Nascimento, 2018a | 60+ | 243 | 316 | Hazard ratio | 1.7 (1.5-1.9); | 2.20 (1.85 - 2.63) |  | Moderate |
| Nascimento, 2022 | 60+ | 255 | not reported | hazard ratio |  | 2.35 (1.64 - 3.39) | 1.80 (1.20 - 2.70) | Low |
| **Psychosocial** |  |  |  |  |  |  |  |  |
| Fekadu, 2015 | 15 - 49 | 121 | Not reported | SMR |  | 2.14 ( 1.77–2.56) |  | low |
| Li, Y 2021 | 15+ | 2,579 | Not reported | SMR |  | 1.50 (1.42-1.58) |  | Low |
| **Visual** |  |  |  |  |  |  |  |  |
| Agrawal, 2011a | 60+ | 15 | 85 | Hazard ratio | 1.8 (1.0-3.1) | 0.81 (0.45-1.47) | 0.75 (0.41-1.38) | Low |
| Ascencio, 2022c | 60+ | 62 | not reported | hazard ratio | 1.05 (0.62 - 1.77) |  |  | high |
| Cao, 2021 | 30+ | 61 | 349 | Hazard ratio |  |  | 0.98 (0.84 - 1.14) | Low |
| Gu, 2013 | 65+ | Not reported | Not reported | Hazard ratio | Women 65-79: 2.8 (1.8-4.2); Women 80+: 1.4 (1.3-1.6); Men 65-79: 2.0 (1.2-3.2); Men 80+: 1.5 (1.4-1.7) |  |  | High |
| Khanna, 2013 | 30+ | 105 | 326 | Hazard ratio | 6.3 (5.0-7.8) |  | 1.9 (1.5-2.5) | Low |
| Kuper, 2019 | >50 | 100 | 309 | Risk ratio |  | 1.54 (1.22 - 1.93) | 1.37 (1.10 - 1.71) | Low |
| Li Z, 2011 | 50-96 | 13 | 161 | Odds ratio |  | 3.4 (2.0-6.6) |  | Moderate |
| Ojagbemi, 2017b | 65+ | Not reported | Not reported | Risk ratio |  |  | 1.7 (0.4-7.4) | Low |
| Pion,2002 | 40+ | 38 (37.6%) | 25 (24.8%) | Risk ratio |  | 2.3 (1.10–4.83) | 2.2 (1.20-4.05) | Moderate |
| Sun, 2020a | 65+ | 4569 | 9128 | Hazard ratio |  | 1.3 (1.2-1.3) | 1.2 (1.2-1.2) | Low |
| Taylor, 1991 | All ages | 12 | 7 | Odds Ratio | 3.0 (1.3-7.2) | 3.33 (1.03-11.04) |  | Low |
| Wang Z, 2022 | 45+ | not reported | not reported | Relative risk | 1.29 (1.17 to 1.44) |  | 1.22 (1.08 to 1.37) | Low |

\*SMR = standardised mortality ratio

Sixty-two cohorts from 53 studies were eligible for inclusion in the meta-analysis, comprising 267,415 participants with disabilities. The remaining studies identified in the systematic review were not included due to reporting measures of effect that could not be pooled with HRs (31, 34, 39, 47, 56, 73, 93, 95), and reporting of unadjusted measures of effect (21, 22, 29, 35, 43, 55, 61, 71, 88).

The pooled maximally adjusted HR for mortality in people with disabilities compared with those without disabilities was 2.06 (95%CI 1.80 - 2.34) and heterogeneity between studies was high (τ^²^=0·24, I^²^=98%) suggesting differing effects across studies (Supplementary file 3). This association varied by impairment type; 1.91 (95%CI: 1.61 – 2.26) for cognitive impairment, 1.95 (1.67 – 2.27) for functional impairment, 1.40 (95%CI 1.30 – 1.51) for hearing impairment, 2.39 (95%CI 1.86 – 3.07) for neurological impairment, 3.39 (0.90 – 12.76) for multiple impairments, 1.77 (95%CI 1.25 – 2.51) for psychosocial impairment and 1.32 (95%CI 1.13 – 1.55) for visual impairment (Figure 2).

**Figure 2:**
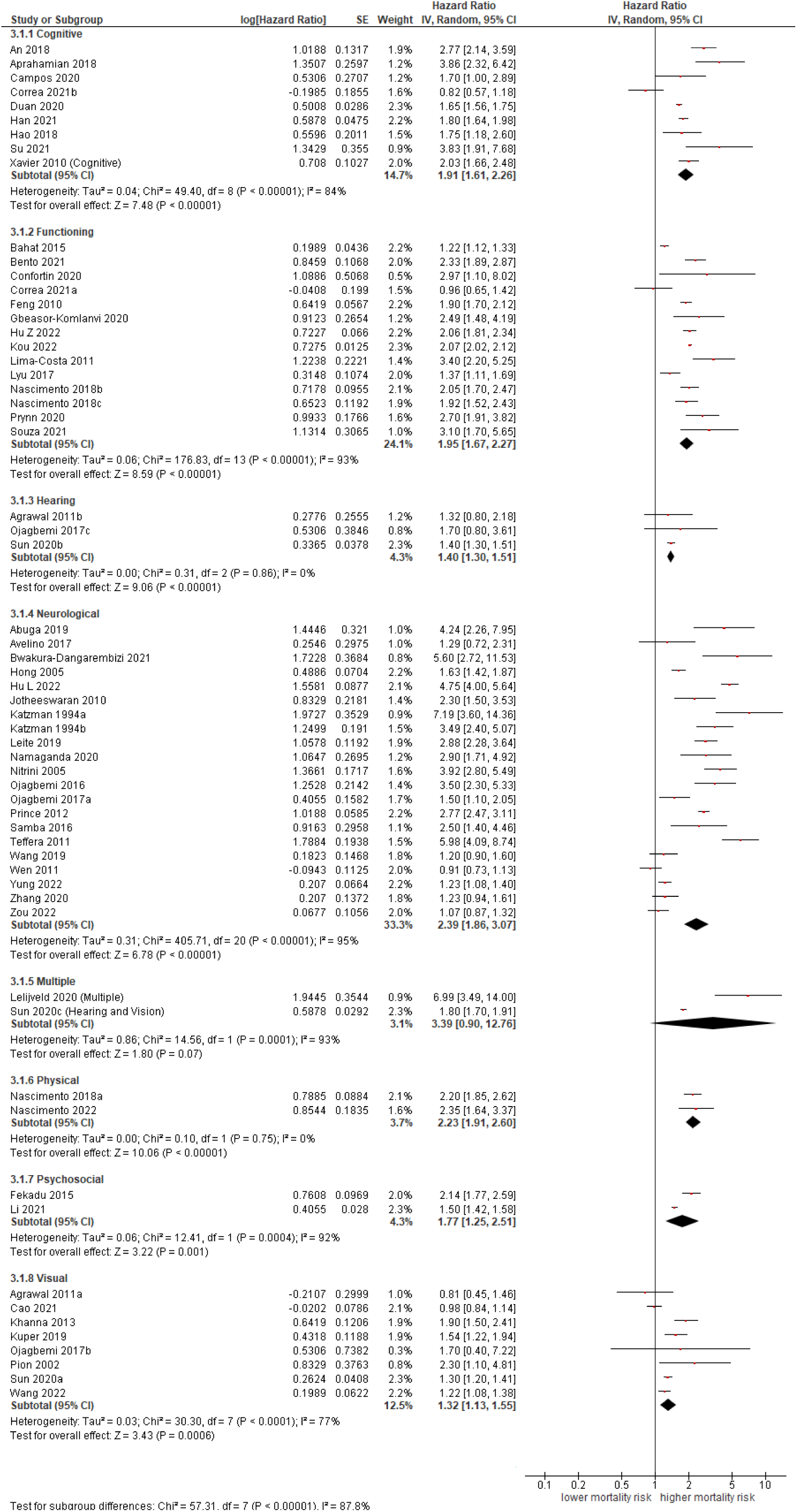
Association of impairment type with all-cause mortality

Four studies compared the hazard for mortality among children with disabilities versus children without disabilities. Pooled HRs for all-cause mortality was 4.46 (95%CI 3.01–6.59) for children under 18 with disabilities versus those without disabilities, with low heterogeneity between studies (τ^²^=0·05, I²=34%) that suggests a consistent effect across studies (Figure 3). Two studies compared the hazard for mortality in participants with disabilities aged 15 – 49 versus those without disabilities, with a HR for mortality of 3·53 (95%CI 1·29–9·66, τ^²^=0·50, I^²^=96%). Twenty-seven studies (35 cohorts) compared the hazard for mortality in participants over the age of 60 with disabilities to those without disabilities; the adjusted HR for mortality was 1·97 (95%CI 1·63–2.38, τ^²^=0·28, I^²^=99%).

**Figure 3:**
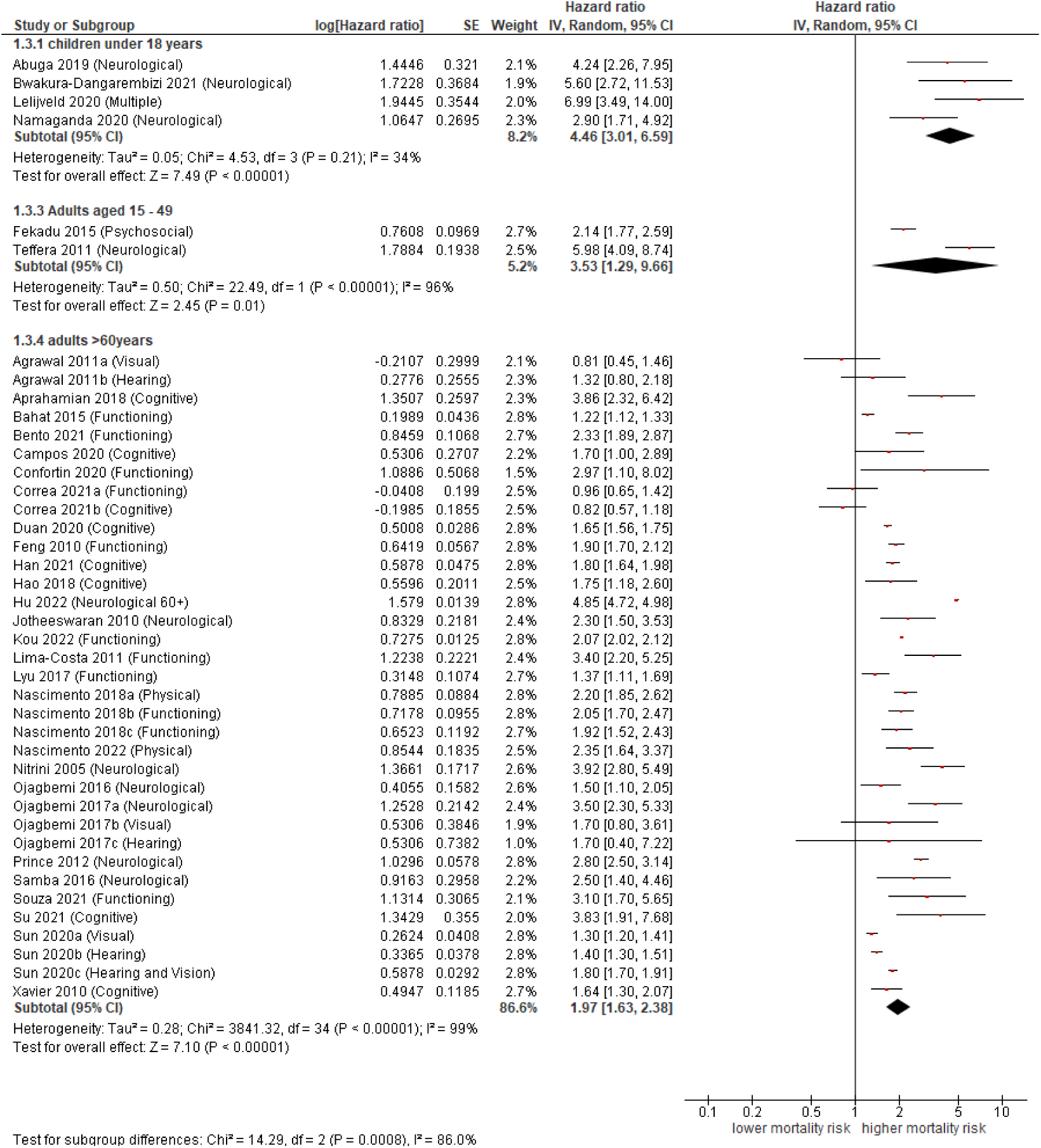
Random effects meta-analysis by age group

Meta-regression analysis of studies comparing all-cause mortality between the participants with disabilities and those without revealed that the association between disability and mortality was not influenced by risk of bias (Chi^2^ 3.03, p=0.22). Funnel plots were reviewed for studies comparing all-cause mortality in participants with disabilities with participants who did not have a disability, and no evidence of publication bias was identified (Supplementary file 4).

## Discussion

### Review of findings

There is limited knowledge about all-cause mortality for people with disability in general, and within LMIC settings specifically. This systematic review and meta-analysis synthesizes the available data from 22 countries and 5 WHO regions on the association between disability and the risk of mortality in LMICs. We found an increased risk of early death for people with disabilities across impairment types and in all ages. The association varied by impairment type as it was strongest for people with multiple impairments, followed by those with neurological impairment. The risk of early death is greatest for children with disabilities and remains increased for adults aged 15 – 49 as well as adults over 60 years of age. All 17 heterogenous studies that were identified in the systematic review and that were not included in the meta-analysis reported an association between disability and increased mortality.

There was little evidence that the quality of the studies or publication bias impacted on the results. Whilst the quality of the included studies was generally high, this review highlighted that there is variability in the methods used to assess and report disability, as well as mortality. Studies predominantly used clinical diagnosis followed by self-report to evaluate disability status and verbal autopsy and population registers to establish death.

### Consistency with other reviews

Our results are consistent with meta-analyses of specific impairments and mortality. Globally, Ehrlich et al (13) found that people with visual impairment have a higher risk of early death compared to those with normal vision and the magnitude of the effect increased with severity of impairment. O’Leary et al (15) established that people with intellectual impairments are missing 20 years of life experience globally, and that age of death is lower for people with more severe intellectual disabilities and for people with additional comorbidities such as epilepsy, genetic syndromes and functional impairments. Abuga et al (96) identified that the risk of early death is increased in children with neurological impairments and adults with child onset neurological impairments compared with the general population (median SMR was 2.9 (range 2.0–11.6)), with few studies eligible for inclusion from LMIC.

### Limitations and strengths

This systematic review and meta-analysis has limitations that need to be taken into account. Whilst we placed no restrictions on language, the electronic searches were conducted in the English language, and some literature may have been missed. Data was missing from the Eastern Mediterranean region, with only four studies representing the South-East Asia and European regions. Therefore, included studies may not be representative of all LMIC. Nevertheless, 62 cohorts from 22 countries were included in the meta-analysis. Included studies demonstrate a significant association between disability and all-cause mortality in both multivariable adjusted and age-adjusted pooled effect estimates. We included multivariable adjusted studies where the age-adjusted estimates were unavailable. These studies adjusted for other important factors that could explain the association between disability and mortality, such as socio-economic status and comorbidities, which are common risk factors for disability and mortality (3–6). They may therefore have over-adjusted the statistical models and included variables that lie on the causal pathway between disability and mortality. This adjustment could have biased study results toward no effect. Underreporting of disability in studies using self-report is a potential concern, as it may lead to misclassification and bias of the association towards the null . There were also inconsistencies in the assessment of mortality, disability and adjustments made in the analyses, which make comparisons difficult and may explain the high heterogeneity in the meta-analyses.

This study has several key strengths. We included studies from regions in the world that are not represented in any other meta-analyses for disability and mortality. We also included a risk of bias assessment for the included studies. The quality of included studies was generally high and nearly three quarters of studies were estimated to have a low risk of bias across all domains. There was no evidence from the meta-regression analyses that the estimated association was affected by risk of bias.

### Implications for research, policy and practice

Our findings that all people with disabilities are at risk of early death in LMIC have important implications for research, policy and practice. Consistent methods of measurement and disaggregation of results by disability are imperative to monitor progress toward the SDGs with respect to disability. Our study highlights the need for widespread adoption of standard definitions and protocols to promote comparability across cohorts. There was notably more information on conditions, such as schizophrenia and dementia, which are frequently recorded in medical records, while other groups (e.g. physical impairment, hearing impairment) were lacking. Attention is therefore needed on how to record disability and impairment type more consistently in medical records to allow use of this resource to measure mortality trends. Research is also needed to develop and test interventions to reduce mortality among people with disabilities, which may include health system strengthening (e.g. healthcare worker training on disability), improving affordability of care for people with disabilities (e.g. provision of health insurance) and tackling the social determinants of poor health in this group.

A key implication of our review is that it will be difficult to reach SDG3, in particular targets related to reducing mortality (e.g. SDG3.2 - “end all preventable deaths under five years of age”), if the one billion people with disabilities globally continue to experience higher mortality. The global community must therefore design disability-inclusive health systems to promote health, wellbeing and rehabilitation for people with disabilities. These systems must “expect, accept and connect” people with disabilities, meaning that they are able to reach accessible services, experience positive attitudes from healthcare workers, and are connected to further services that they need (97). Services for people with disabilities should be included in national health systems at all levels to maximise access and targeted healthcare improvements required for people with disabilities are available when needed (e.g. health promotion programs that focus on lifestyle behaviours and wellness (98)). Reasonable accommodation must be made within provision of quality health care. For example, for people with intellectual impairment, inclusion of anticipatory care in the form of health checks has been shown to improve the management of long-term conditions and quality of life, in addition to having greater clinical and cost-effectiveness when compared with standard care (99, 100).

## Conclusion

Disability increases the risk of all-cause mortality in LMICs, particularly in childhood. Interventions are needed to improve health of people with disabilities and reduce their risk of death. Without a focus on disability, it may be difficult to reach SDG3 and other key global health targets.

### Contributors

TS and HK conceived the study, developed the research protocol and all data collection instruments and co-led data collection. TS led analysis. TS wrote the first draft of the manuscript with substantial input from HK. Both authors approved of the final version for publication.

### Funding

This project is funded by the National Institute for Health Research (NIHR) [NIHR Global Research Professorship (Grant Reference Number NIHR301621)] awarded to Prof. Hannah Kuper. Hannah Kuper has also received support from the Programme for Evidence to Inform Disability Action (PENDA) grant from FCDO.

### Disclaimer

The views expressed in this publication are those of the author(s) and not necessarily those of the NIHR, NHS, the UK Department of Health and Social Care or FCDO.

### Patient consent for publication

Not required.

### Provenance and peer review

Not commissioned; externally peer reviewed.

### Data sharing statement

No additional data are available

## Data Availability

All data produced in the present work are contained in the manuscript

---

**Supplementary file 1:**
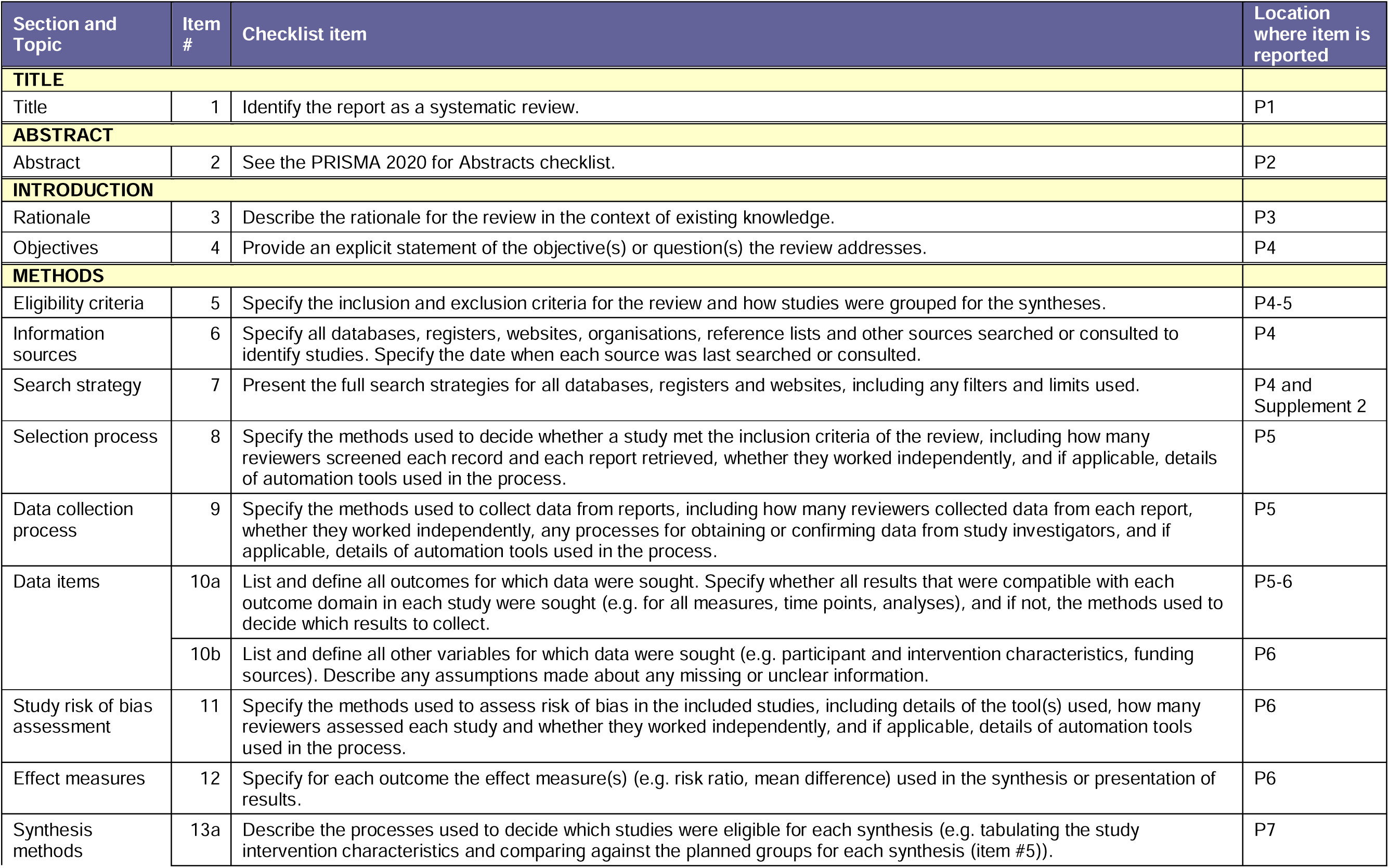
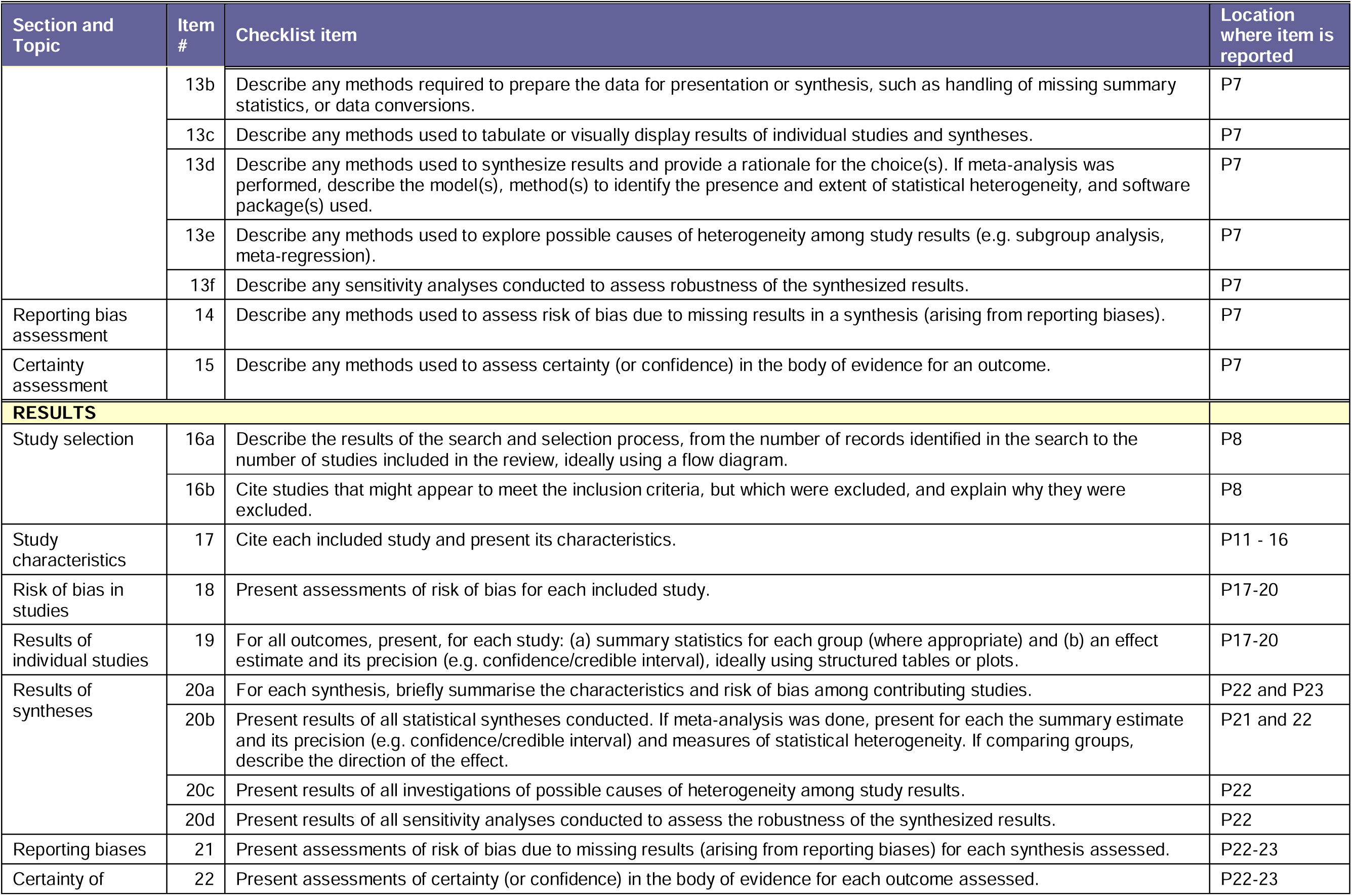
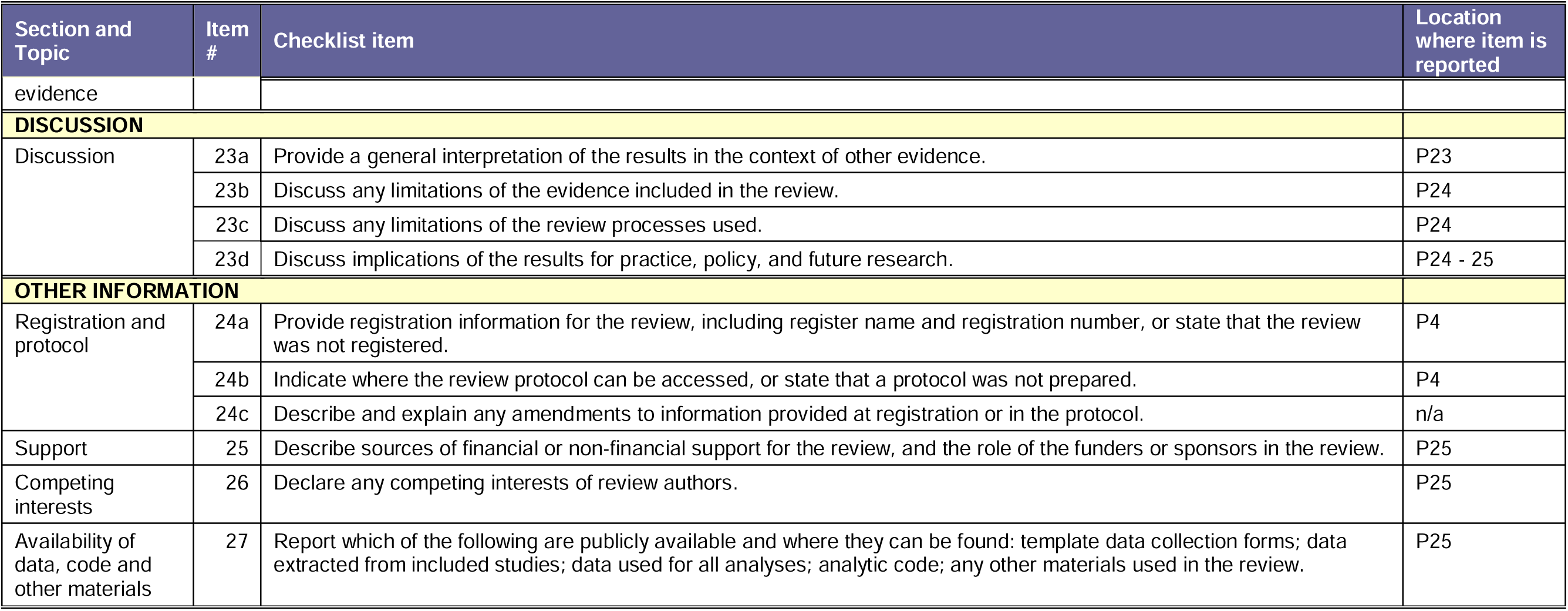
Preferred Reporting Items for Systematic Reviews and Meta-Analyses guidelines

| Section and Topic | Item # | Checklist item | Location where item is reported |
| --- | --- | --- | --- |
| **TITLE** |  |  |  |
| Title | 1 | Identify the report as a systematic review. | P1 |
| **ABSTRACT** |  |  |  |
| Abstract | 2 | See the PRISMA 2020 for Abstracts checklist. | P2 |
| **INTRODUCTION** |  |  |  |
| Rationale | 3 | Describe the rationale for the review in the context of existing knowledge. | P3 |
| Objectives | 4 | Provide an explicit statement of the objective(s) or question(s) the review addresses. | P4 |
| **METHODS** |  |  |  |
| Eligibility criteria | 5 | Specify the inclusion and exclusion criteria for the review and how studies were grouped for the syntheses. | P4-5 |
| Information sources | 6 | Specify all databases, registers, websites, organisations, reference lists and other sources searched or consulted to identify studies. Specify the date when each source was last searched or consulted. | P4 |
| Search strategy | 7 | Present the full search strategies for all databases, registers and websites, including any filters and limits used. | P4 and Supplement 2 |
| Selection process | 8 | Specify the methods used to decide whether a study met the inclusion criteria of the review, including how many reviewers screened each record and each report retrieved, whether they worked independently, and if applicable, details of automation tools used in the process. | P5 |
| Data collection process | 9 | Specify the methods used to collect data from reports, including how many reviewers collected data from each report, whether they worked independently, any processes for obtaining or confirming data from study investigators, and if applicable, details of automation tools used in the process. | P5 |
| Data items | 10a | List and define all outcomes for which data were sought. Specify whether all results that were compatible with each outcome domain in each study were sought (e.g. for all measures, time points, analyses), and if not, the methods used to decide which results to collect. | P5-6 |
|  | 10b | List and define all other variables for which data were sought (e.g. participant and intervention characteristics, funding sources). Describe any assumptions made about any missing or unclear information. | P6 |
| Study risk of bias assessment | 11 | Specify the methods used to assess risk of bias in the included studies, including details of the tool(s) used, how many reviewers assessed each study and whether they worked independently, and if applicable, details of automation tools used in the process. | P6 |
| Effect measures | 12 | Specify for each outcome the effect measure(s) (e.g. risk ratio, mean difference) used in the synthesis or presentation of results. | P6 |
| Synthesis methods | 13a | Describe the processes used to decide which studies were eligible for each synthesis (e.g. tabulating the study intervention characteristics and comparing against the planned groups for each synthesis (item #5)). | P7 |
|  | 13b | Describe any methods required to prepare the data for presentation or synthesis, such as handling of missing summary statistics, or data conversions. | P7 |
|  | 13c | Describe any methods used to tabulate or visually display results of individual studies and syntheses. | P7 |
|  | 13d | Describe any methods used to synthesize results and provide a rationale for the choice(s). If meta-analysis was performed, describe the model(s), method(s) to identify the presence and extent of statistical heterogeneity, and software package(s) used. | P7 |
|  | 13e | Describe any methods used to explore possible causes of heterogeneity among study results (e.g. subgroup analysis, meta-regression). | P7 |
|  | 13f | Describe any sensitivity analyses conducted to assess robustness of the synthesized results. | P7 |
| Reporting bias assessment | 14 | Describe any methods used to assess risk of bias due to missing results in a synthesis (arising from reporting biases). | P7 |
| Certainty assessment | 15 | Describe any methods used to assess certainty (or confidence) in the body of evidence for an outcome. | P7 |
| **RESULTS** |  |  |  |
| Study selection | 16a | Describe the results of the search and selection process, from the number of records identified in the search to the number of studies included in the review, ideally using a flow diagram. | P8 |
|  | 16b | Cite studies that might appear to meet the inclusion criteria, but which were excluded, and explain why they were excluded. | P8 |
| Study characteristics | 17 | Cite each included study and present its characteristics. | P11 - 16 |
| Risk of bias in studies | 18 | Present assessments of risk of bias for each included study. | P17-20 |
| Results of individual studies | 19 | For all outcomes, present, for each study: (a) summary statistics for each group (where appropriate) and (b) an effect estimate and its precision (e.g. confidence/credible interval), ideally using structured tables or plots. | P17-20 |
| Results of syntheses | 20a | For each synthesis, briefly summarise the characteristics and risk of bias among contributing studies. | P22 and P23 |
|  | 20b | Present results of all statistical syntheses conducted. If meta-analysis was done, present for each the summary estimate and its precision (e.g. confidence/credible interval) and measures of statistical heterogeneity. If comparing groups, describe the direction of the effect. | P21 and 22 |
|  | 20c | Present results of all investigations of possible causes of heterogeneity among study results. | P22 |
|  | 20d | Present results of all sensitivity analyses conducted to assess the robustness of the synthesized results. | P22 |
| Reporting biases | 21 | Present assessments of risk of bias due to missing results (arising from reporting biases) for each synthesis assessed. | P22-23 |
| Certainty of | 22 | Present assessments of certainty (or confidence) in the body of evidence for each outcome assessed. | P22-23 |
| evidence |  |  |  |
| **DISCUSSION** |  |  |  |
| Discussion | 23a | Provide a general interpretation of the results in the context of other evidence. | P23 |
|  | 23b | Discuss any limitations of the evidence included in the review. | P24 |
|  | 23c | Discuss any limitations of the review processes used. | P24 |
|  | 23d | Discuss implications of the results for practice, policy, and future research. | P24 - 25 |
| **OTHER INFORMATION** |  |  |  |
| Registration and protocol | 24a | Provide registration information for the review, including register name and registration number, or state that the review was not registered. | P4 |
|  | 24b | Indicate where the review protocol can be accessed, or state that a protocol was not prepared. | P4 |
|  | 24c | Describe and explain any amendments to information provided at registration or in the protocol. | n/a |
| Support | 25 | Describe sources of financial or non-financial support for the review, and the role of the funders or sponsors in the review. | P25 |
| Competing interests | 26 | Declare any competing interests of review authors. | P25 |
| Availability of data, code and other materials | 27 | Report which of the following are publicly available and where they can be found: template data collection forms; data extracted from included studies; data used for all analyses; analytic code; any other materials used in the review. | P25 |

**Supplementary file 2:** Search term example

EMBASE search strategy (14.11.2022)
| # |  | Searches | Results |
| --- | --- | --- | --- |
| 1 | LMIC | Embase expert search with World Bank LMIC from June 2019 | 1,982,116 |
| 2 | Disability | ((person* with disabilit* or people with disabilit* or (disable* or Disabilit* or Handicap*)) adj2 (person* or people or individ* or patient* or subject* or adult* or elderly)).ti,ab | 39,154 |
| 3 |  | (Physical* adj2 (impair* or deficienc* or disable* or disabili* or handicap*)).ti,ab. | 23,216 |
| 4 |  | (Cerebral pals* or Spina bifida or Muscular dystroph* or Osteogenesis imperfecta or Musculoskeletal abnormalit* or Musculo-skeletal abnormalit* or Muscular abnormalit* or Skeletal abnormalit* or Limb abnormalit* or Amputation* or Clubfoot or Poliomyeliti* or Paraplegi* or Paralys* or Paralyz* or Hemiplegi* or wheelchair user* or wheel chair user*).ti,ab. | 297,217 |
| 5 | | ((Hearing or Acoustic or Ear\$3) adj2 (loss* or impair* or deficienc* or disable* or disabili* or handicap*)).ti,ab. | 98,955 |
| 6 | | ((Visual* or Vision or Eye\$3) adj2 (loss* or impair* or deficienc* or disable* or disabili* or handicap*)).ti,ab | 72,364 |
| 7 |  | (Deaf* or Blind*).ti,ab. | 520,496 |
| 8 |  | exp Hearing impairment/ or exp vision disorders/ or exp Deafness/ or exp Blindness/ | 388,253 |
| 9 |  | (Schizophreni* or Psychosis or Psychoses or Psychotic Disorder* or Schizoaffective Disorder* or Schizophreniform Disorder* or Dementia* or Alzheimer*).ti,ab. | 557,897 |
| 10 |  | exp Schizophrenia/ | 206,296 |
| 11 |  | exp Dementia/ or exp Alzheimer disease/ | 400,715 |
| 12 |  | ((Intellectual* or Mental* or Psychological* or Developmental) adj2 (impair* or retard* or deficienc* or disable* or disabili* or handicap* or ill?6)).ti,ab | 104,985 |
| 13 |  | (communication or language or speech or learning) adj2 disorder*).ti,ab. | 14,280 |
| 14 |  | exp Disabled Children/ | 10,005 |
| 15 |  | exp Learning Disorders/ | 39,920 |
| 16 |  | exp Mentally Disabled Persons/ | 1,343 |
| 17 |  | ((genetic or hereditary or inherited or congenital) adj2 (disease* or ill* or syndrome or defect* or disorder* or condition* or malformation or anomal* or abnormalit*)).ti,ab | 306,393 |
| 18 |  | (elderly or older adult*).ti,ab. | 489,760 |
| 19 |  | Or /2 – 18 [ALL DISABILITY TERMS] | 2,720,549 |
| 20 | Mortality | Exp Mortality/ | 1,272,321 |
| 21 |  | Death/ | 324,521 |
| 22 |  | Death Certificate | 9,978 |
| 23 |  | Mortality.tw. | 1,298,152 |
| 24 | | Death\$.tw. | 1,352,807 |
| 25 |  | (Fatality or fatalities).tw | 42,777 |
| 26 |  | Or/ 20 – 25 [ALL MORTALITY TERMS] | 2,742,787 |
| 27 | Study type | Cohort analysis/ | 781,100 |
| 28 |  | Longitudinal study/ | 165,118 |
| 29 | | (Cohort\$ or longitudinal).tw | 1,510,634 |
| 30 |  | Cross-sectional study/ | 447,938 |
| 31 |  | questionnaire/ | 787,233 |
| 32 |  | Health survey/ or health care survey/ | 231,915 |
| 33 |  | (Survey or surveys).tw | 907,884 |
| 34 |  | Or/ 27 – 33 [SPECIFIC STUDY DESIGNS] | 3,368,663 |
| 35 |  | 1 AND 19 AND 26 AND 34 | 5,134 |
| 36 |  | Limit to >1990, humans | 5,061 |

**Supplementary file 3:**
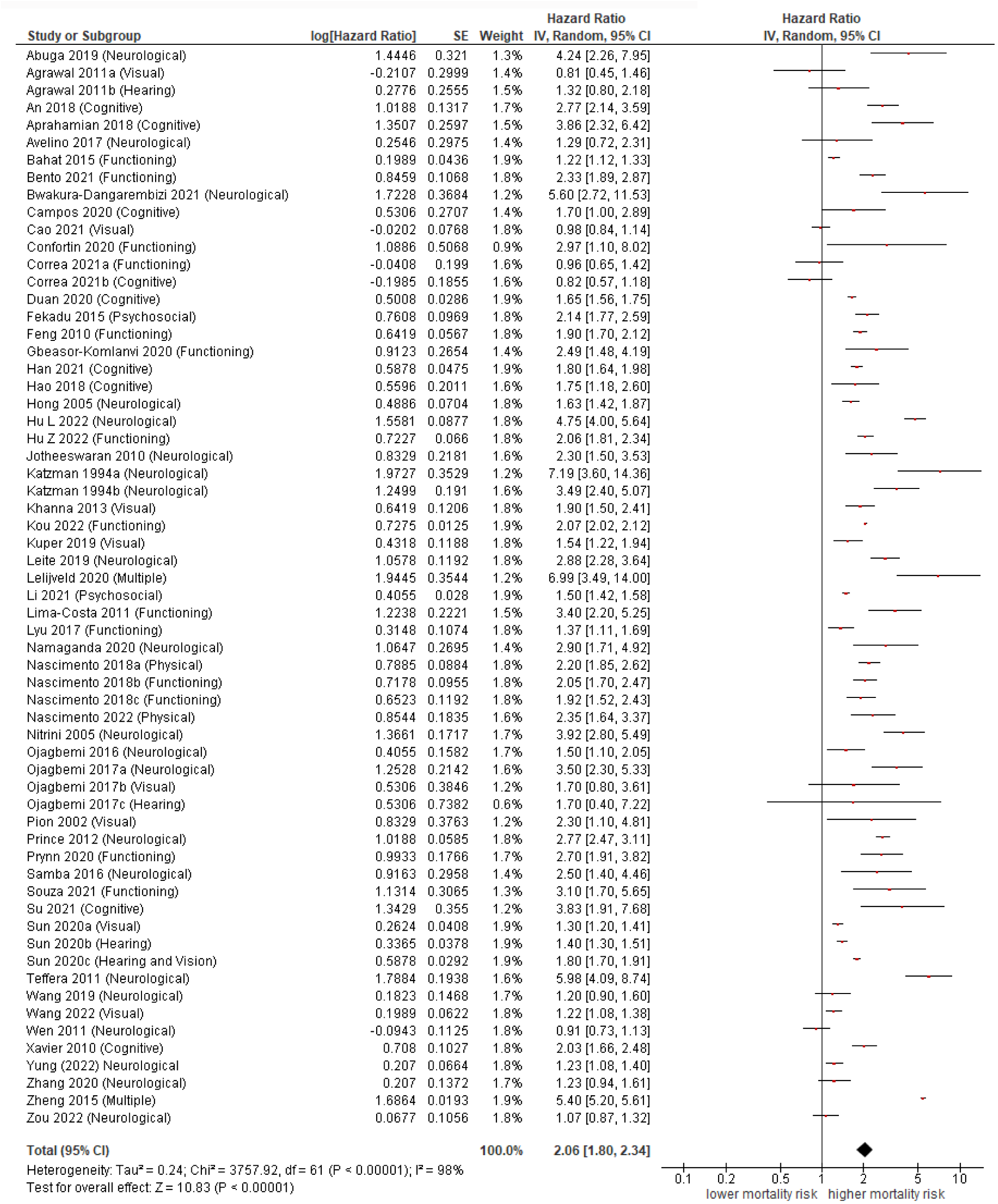
Association of disability with all-cause mortality

**Supplementary file 4:**
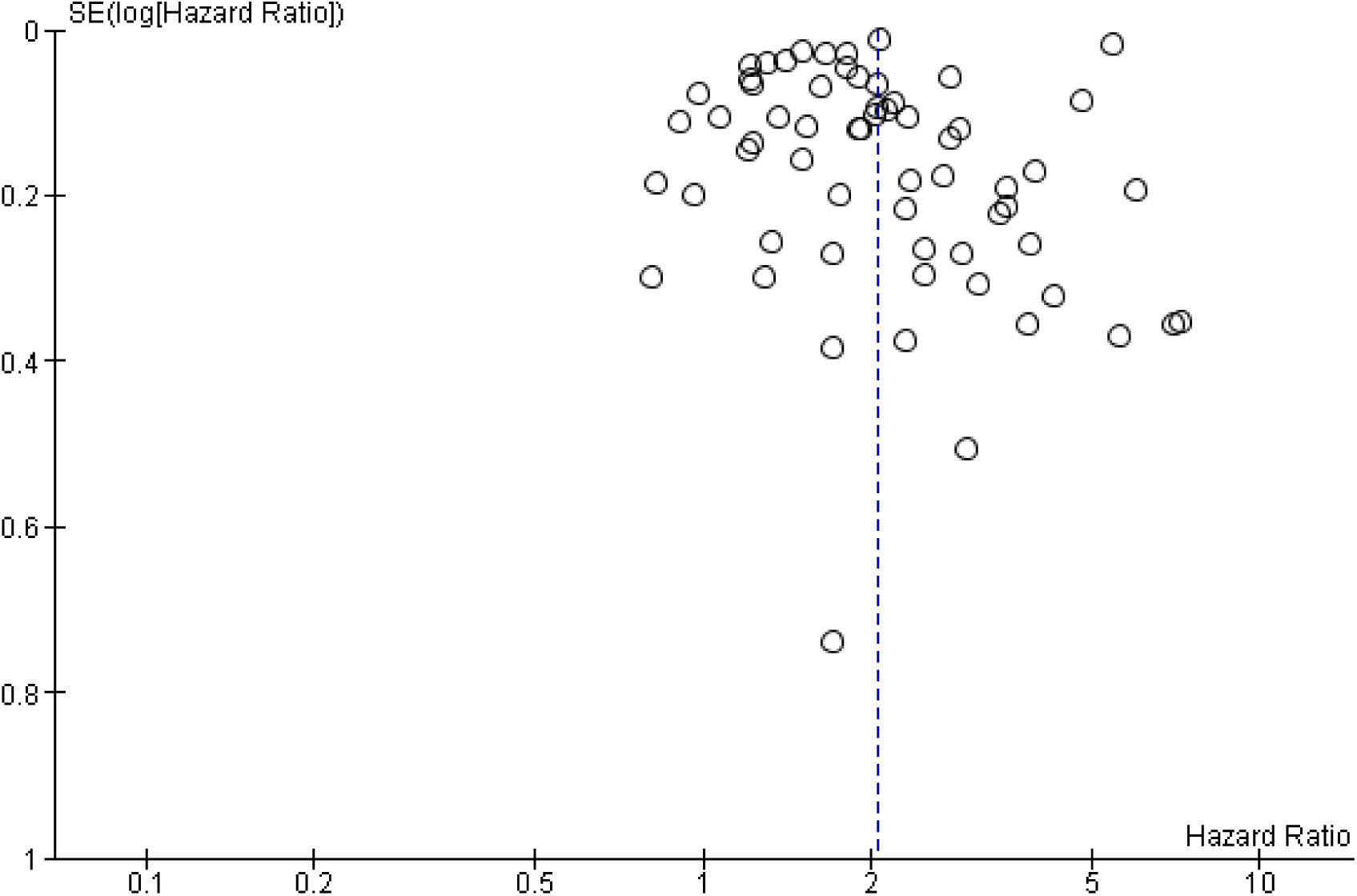
Funnel plot for studies assessing all-cause mortality in people with disabilities

